# Cross-national factors associated with governmental responses to the 2025 PEPFAR funding freeze

**DOI:** 10.64898/2026.10.18.26365861

**Authors:** Andrew McGregor, Sahan Suggala, Wesam Abdeljaber, Jessica Michael, Brittany Shoemake

## Abstract

Since its inception in 2003, the United States President’s Emergency Plan for AIDS Relief (PEPFAR) has played a central role in HIV/AIDS prevention and treatment across Sub-Saharan Africa. Following the January 2025 suspension of United States foreign aid, which includes PEPFAR funding, countries across Sub-Saharan Africa demonstrated varying governmental responses to the resulting HIV funding disruptions. This study evaluates whether national characteristics were associated with a country’s ability to respond to this funding crisis. Countries in Sub-Saharan Africa receiving PEPFAR funding were classified into one of three response categories: Level 1 (minimal or no governmental response), Level 2 (government response without sufficient plans or resources to fully offset funding losses), or Level 3 (high governmental response with strategies to maintain HIV/AIDS prevention and treatment services). Response categories were compared with national Human Development Index (HDI), GDP per capita, HIV prevalence, total population living with HIV, annual HIV mortality, and historical PEPFAR funding. Univariable proportional-odds ordinal logistic regression was used to identify factors associated with response level.

Five countries (17.2%) were classified as Level 1, 10 (34.5%) as Level 2, and 14 (48.3%) as Level 3. Higher HDI was significantly associated with stronger governmental response (OR = 4.05; 95% CI, 1.49–11.04; Holm-adjusted p = 0.0376). GDP per capita (OR = 2.35; 95% CI, 1.00–5.51; unadjusted p = 0.0489) and HIV prevalence (OR = 3.53; 95% CI, 1.03–12.11; unadjusted p = 0.0452) demonstrated positive associations before adjustment for multiple comparisons, but neither remained significant after Holm correction (adjusted p = 0.2261 for both). Historical PEPFAR funding, total population living with HIV, and annual HIV mortality were not significantly associated with response level after adjustment.

Countries with higher levels of human development demonstrated stronger governmental responses to the 2025 PEPFAR funding disruptions. Several countries with minimal documented responses were also affected by active conflict or political instability, potentially compounding their vulnerability to treatment interruptions. These findings suggest that broader national development may play an important role in governmental responses to funding disruptions and highlight the need to strengthen resilient health systems, diversify HIV financing, and prioritize support for countries at greatest risk of future funding shocks.

## Introduction

Since its inception in 2003, the United States President’s Emergency Plan for AIDS Relief (PEPFAR) has served as a cornerstone of HIV/AIDS prevention and treatment efforts across Sub-Saharan Africa and has represented the single largest contributor to HIV/AIDS funding in the region.**[1, 2]** In January 2025, United States President Donald Trump unexpectedly halted foreign aid payments, including funding mechanisms supporting PEPFAR programs.**[3–6]**

In response to these funding disruptions, governments across Sub-Saharan Africa demonstrated varying levels of preparedness and policy response. Some governments proposed replacement funding strategies or restructuring plans to maintain continuity of HIV/AIDS care delivery, while others demonstrated limited or no publicly articulated response.

Several modelling studies released following the funding interruptions projected substantial increases in HIV transmission and mortality depending on national disease burden, treatment coverage, and reliance on external aid.**[3, 4, 7]** Despite differences in methodology and modelling platforms, these studies consistently demonstrated that even temporary disruptions in antiretroviral therapy delivery networks can produce prolonged downstream effects.**[8, 9]** Medication supply chains are fragile, antiretroviral medications remain costly, and interruptions in external funding may result in workforce attrition, treatment discontinuity, and delayed recovery of care infrastructure.**[9–11]**

This study aims to evaluate whether underlying national-level factors were associated with a country’s ability to respond to a major HIV/AIDS funding crisis. Because HIV/AIDS mortality had declined substantially across the region in part due to sustained PEPFAR support,**[12, 13]** understanding which countries demonstrated stronger adaptive responses may help identify areas where future investments in resilient HIV care delivery systems and domestic program sustainability should be prioritized.**[4]**

## Methods

### Data Collection

Official government announcements, Ministry of Health publications, press releases, and third-party news reports were identified through a targeted search engine–based review between January 2025 and April 2026. Twenty-nine sub-Saharan African country classifications were independently evaluated by three investigators using the predefined framework, while countries with disagreement between reviewers or uncertainty regarding their final classification were considered for additional assessment. A fourth reviewer independently evaluated six selected countries (Benin, Burundi, Eswatini, Kenya, Lesotho, and Liberia) using the same classification criteria. Any disagreements were resolved through discussion until consensus was reached. Responses were evaluated according to three domains:

1. Government acknowledgment of the funding disruption.
2. Specific mitigation strategies (e.g., emergency funding, procurement plans, ART continuity measures).
3. Demonstrated domestic resource mobilization or implementation plans.

**Level 1 (Minimal Response)** was assigned to countries that demonstrated little or no publicly documented governmental response. These countries met one or more of the following criteria:

1. No meaningful public response
2. No replacement funding announced
3. No mitigation measures announced

**Level 2 (Moderate Response)** was assigned to countries that publicly acknowledged the funding disruption and announced partial mitigation measures but did not demonstrate sufficient commitments to fully address the anticipated funding gap. These countries met one or more of the following criteria:

1. Public government acknowledgement of the funding disruption
2. At least partial mitigation measures announced
3. The funding gap remains only partially addressed

**Level 3 (High Response)** was assigned to countries demonstrating substantial governmental action to maintain HIV/AIDS services. These countries met one or more of the following criteria:

1. Significant domestic funding committed
2. Announced and detailed implementation plan
3. Explicit measures to maintain ART and other HIV services

Several countries demonstrated borderline characteristics between response categories. Sensitivity analyses were performed by restoring the prior classifications of five countries (Burundi, Lesotho, Liberia, South Africa, and Tanzania), individually and collectively, and by excluding all five countries from the analysis. Individual reviewer ratings, fourth-reviewer assessments, final consensus classifications, and supporting evidence are provided in **S1 Appendix**.

Country-level HIV prevalence, total population living with HIV, and annual AIDS-related mortality estimates for 2024 were obtained from UNAIDS. Human Development Index (HDI) values were obtained from the United Nations Development Programme’s 2025 Human Development Report (2023 estimates). Nominal GDP per capita values were based on 2026 projections from the International Monetary Fund’s April 2026 World Economic Outlook database. Fiscal year 2024 PEPFAR funding allocations were obtained from publicly available PEPFAR Country Operational Plan data.

### Calculations

Statistical analyses were performed using R (Version 4.3.3). Country-level Human Development Index (HDI), Gross Domestic Product (GDP) per capita, HIV prevalence among persons aged 15–49 years (UNAIDS estimate), total population living with HIV, annual HIV mortality, and historical PEPFAR funding were evaluated. Response category (Levels 1–3) was treated as an ordered categorical dependent variable. Each factor was examined separately using univariable proportional-odds ordinal logistic regression. GDP per capita, total population living with HIV, HIV mortality, and PEPFAR funding were natural-log transformed because of their right-skewed distributions and allowed multiplicative differences to be represented more appropriately. Predictors were standardized so that ordinal odds ratios represented the change in the cumulative odds of membership in a higher response category associated with a one-standard-deviation increase in each factor. Odds ratios, 95% confidence intervals, and two-sided p-values were reported. Because six primary associations were tested, Holm-adjusted p-values were calculated to control the family-wise error rate, with statistical significance defined as an adjusted p-value below 0.05. Separate univariable linear regressions treating response category as a numeric variable from 1 to 3 were performed as sensitivity analyses. An additional sensitivity analysis was conducted using 2024 World Bank GDP per capita estimates in current US dollars. Because predictors were examined separately, multicollinearity diagnostics were not relevant to these univariable models.

### Ethics Statement

This study exclusively used publicly available, country-level data and government reports. No identifiable individual-level data were collected, and no human participants were directly involved.

## Results

Twenty-nine PEPFAR-supported countries in Sub-Saharan Africa were included in the analysis. Five countries (17.2%) were classified as Level 1, 10 (34.5%) as Level 2, and 14 (48.3%) as Level 3. Country-level characteristics and governmental response classifications are in Table 1. While **Fig 1** presents the map of the country-level results.

**Table 1.**
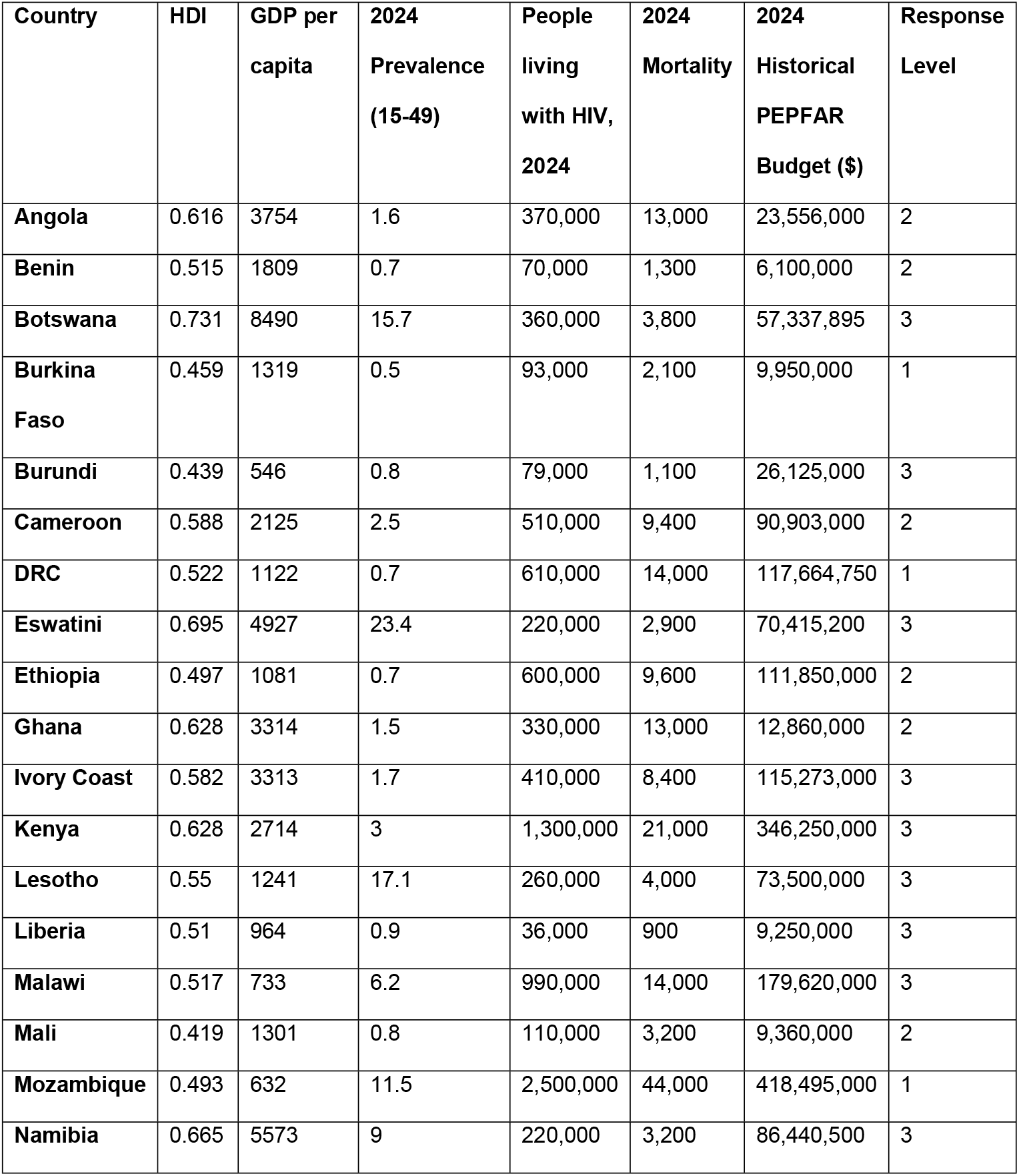
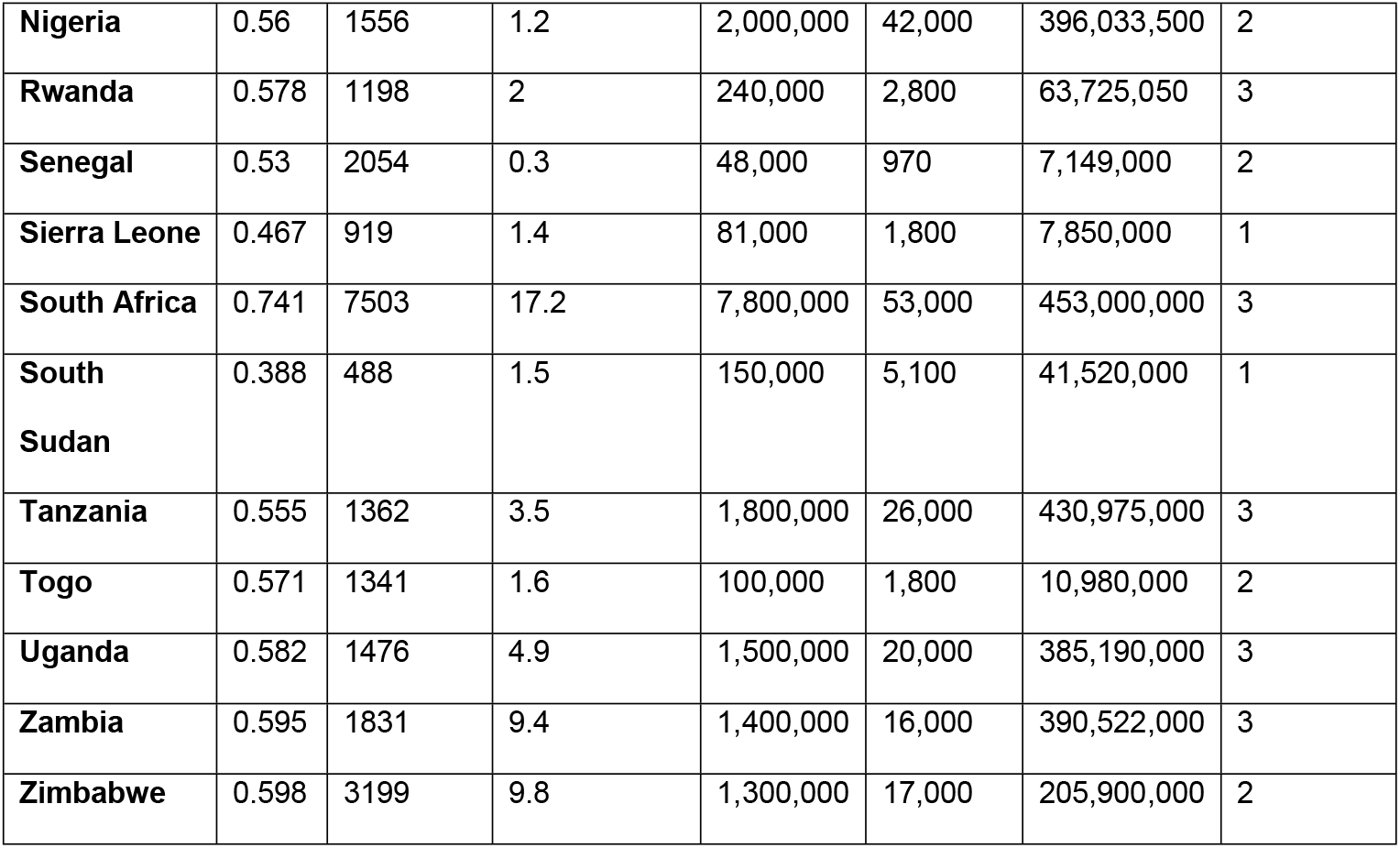
Study variables and governmental response classifications for 29 PEPFAR-supported countries in Sub-Saharan Africa.

| Country | HDI | GDP per capita | 2024 Prevalence (15-49) | People living with HIV, 2024 | 2024 Mortality | 2024 Historical PEPFAR Budget (\$) | Response Level |
| --- | --- | --- | --- | --- | --- | --- | --- |
| Angola | 0.616 | 3754 | 1.6 | 370,000 | 13,000 | 23,556,000 | 2 |
| Benin | 0.515 | 1809 | 0.7 | 70,000 | 1,300 | 6,100,000 | 2 |
| Botswana | 0.731 | 8490 | 15.7 | 360,000 | 3,800 | 57,337,895 | 3 |
| Burkina Faso | 0.459 | 1319 | 0.5 | 93,000 | 2,100 | 9,950,000 | 1 |
| Burundi | 0.439 | 546 | 0.8 | 79,000 | 1,100 | 26,125,000 | 3 |
| Cameroon | 0.588 | 2125 | 2.5 | 510,000 | 9,400 | 90,903,000 | 2 |
| DRC | 0.522 | 1122 | 0.7 | 610,000 | 14,000 | 117,664,750 | 1 |
| Eswatini | 0.695 | 4927 | 23.4 | 220,000 | 2,900 | 70,415,200 | 3 |
| Ethiopia | 0.497 | 1081 | 0.7 | 600,000 | 9,600 | 111,850,000 | 2 |
| Ghana | 0.628 | 3314 | 1.5 | 330,000 | 13,000 | 12,860,000 | 2 |
| Ivory Coast | 0.582 | 3313 | 1.7 | 410,000 | 8,400 | 115,273,000 | 3 |
| Kenya | 0.628 | 2714 | 3 | 1,300,000 | 21,000 | 346,250,000 | 3 |
| Lesotho | 0.55 | 1241 | 17.1 | 260,000 | 4,000 | 73,500,000 | 3 |
| Liberia | 0.51 | 964 | 0.9 | 36,000 | 900 | 9,250,000 | 3 |
| Malawi | 0.517 | 733 | 6.2 | 990,000 | 14,000 | 179,620,000 | 3 |
| Mali | 0.419 | 1301 | 0.8 | 110,000 | 3,200 | 9,360,000 | 2 |
| Mozambique | 0.493 | 632 | 11.5 | 2,500,000 | 44,000 | 418,495,000 | 1 |
| Namibia | 0.665 | 5573 | 9 | 220,000 | 3,200 | 86,440,500 | 3 |
| **Nigeria** | 0.56 | 1556 | 1.2 | 2,000,000 | 42,000 | 396,033,500 | 2 |
| **Rwanda** | 0.578 | 1198 | 2 | 240,000 | 2,800 | 63,725,050 | 3 |
| **Senegal** | 0.53 | 2054 | 0.3 | 48,000 | 970 | 7,149,000 | 2 |
| **Sierra Leone** | 0.467 | 919 | 1.4 | 81,000 | 1,800 | 7,850,000 | 1 |
| **South Africa** | 0.741 | 7503 | 17.2 | 7,800,000 | 53,000 | 453,000,000 | 3 |
| **South Sudan** | 0.388 | 488 | 1.5 | 150,000 | 5,100 | 41,520,000 | 1 |
| **Tanzania** | 0.555 | 1362 | 3.5 | 1,800,000 | 26,000 | 430,975,000 | 3 |
| **Togo** | 0.571 | 1341 | 1.6 | 100,000 | 1,800 | 10,980,000 | 2 |
| **Uganda** | 0.582 | 1476 | 4.9 | 1,500,000 | 20,000 | 385,190,000 | 3 |
| **Zambia** | 0.595 | 1831 | 9.4 | 1,400,000 | 16,000 | 390,522,000 | 3 |
| **Zimbabwe** | 0.598 | 3199 | 9.8 | 1,300,000 | 17,000 | 205,900,000 | 2 |

**Fig 1.**
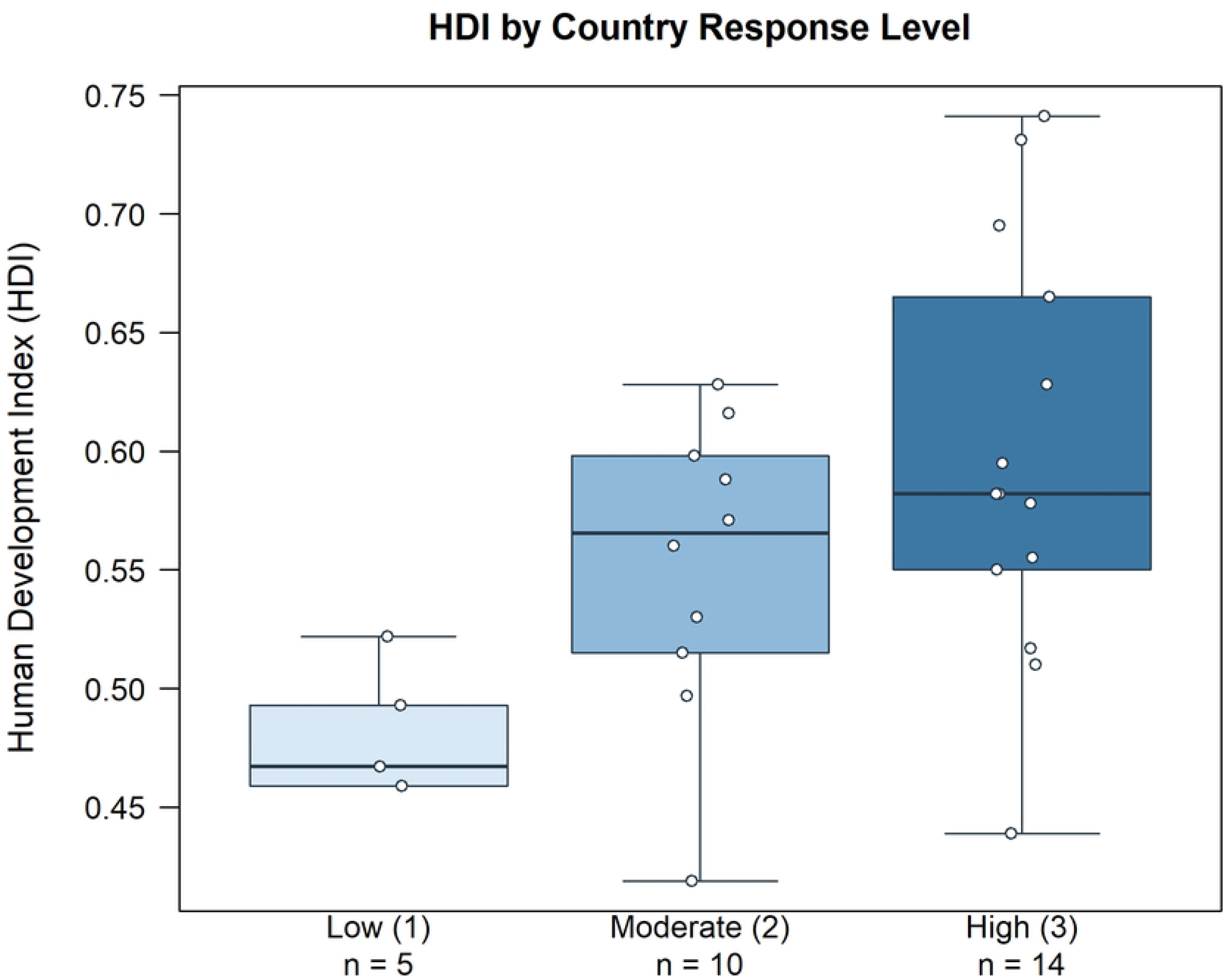
Governmental response classifications across 29 PEPFAR-supported countries in Sub-Saharan Africa.

HDI was significantly and positively associated with country response level in the univariable ordinal regression analysis (OR = 4.05; 95% CI, 1.49–11.04; Holm-adjusted p = 0.0376) (Fig 2). GDP per capita was positively associated with response level before adjustment for multiple comparisons (OR = 2.35; 95% CI, 1.00–5.51; unadjusted p = 0.0489), but the association did not remain significant after Holm correction (adjusted p = 0.2261) (Fig 3). HIV prevalence was also positively associated with response level before adjustment (OR = 3.53; 95% CI, 1.03–12.11; unadjusted p = 0.0452), but not after Holm correction (adjusted p = 0.2261). Historical PEPFAR funding was not significantly associated with response level before or after adjustment (OR = 2.05; 95% CI, 0.97–4.32; unadjusted p = 0.0593; Holm-adjusted p = 0.2261). Total persons living with HIV/AIDS (OR = 1.49; 95% CI, 0.72–3.12; unadjusted p = 0.2847; Holm-adjusted p = 0.5695) and HIV mortality (OR = 1.05; 95% CI, 0.52–2.11; unadjusted and Holm-adjusted p = 0.8963) were also not significantly associated with response category. Thus, HDI was the only country-level factor that remained significantly associated with a stronger governmental response after correction for multiple comparisons. These results are in **Table 2**. Although GDP per capita, HIV prevalence, and historical PEPFAR funding showed positive associations, neither remained statistically significant after adjustment. Because predictors were examined separately, multicollinearity diagnostics were not relevant to these univariable models.

**Table 2.** Associations between national characteristics and governmental response level.

| Factor | Ordinal OR (95% CI) | Ordinal p | Ordinal Holm p | Linear $\beta$ (95% CI) | Linear p | Linear Holm p |
| --- | --- | --- | --- | --- | --- | --- |
| HDI | 4.05 (1.49–11.04) | 0.0063 | 0.0376 | 0.416 (0.164–0.667) | 0.0022 | 0.0129 |
| GDP per capita, log | 2.35 (1.00–5.51) | 0.0489 | 0.2261 | 0.296 (0.020–0.573) | 0.0367 | 0.1808 |
| HIV prevalence | 3.53 (1.03–12.11) | 0.0452 | 0.2261 | 0.297 (0.021–0.574) | 0.0362 | 0.1808 |
| Total persons living with HIV, log | 1.49 (0.72–3.12) | 0.2847 | 0.5695 | 0.143 (–0.152–0.438) | 0.3297 | 0.6593 |
| HIV mortality, log | 1.05 (0.52–2.11) | 0.8963 | 0.8963 | 0.016 (–0.285–0.316) | 0.9160 | 0.9160 |
| Historical PEPFAR funding, log | 2.05 (0.97–4.32) | 0.0593 | 0.2261 | 0.253 (–0.031–0.536) | 0.0783 | 0.2348 |

**Fig 2.**
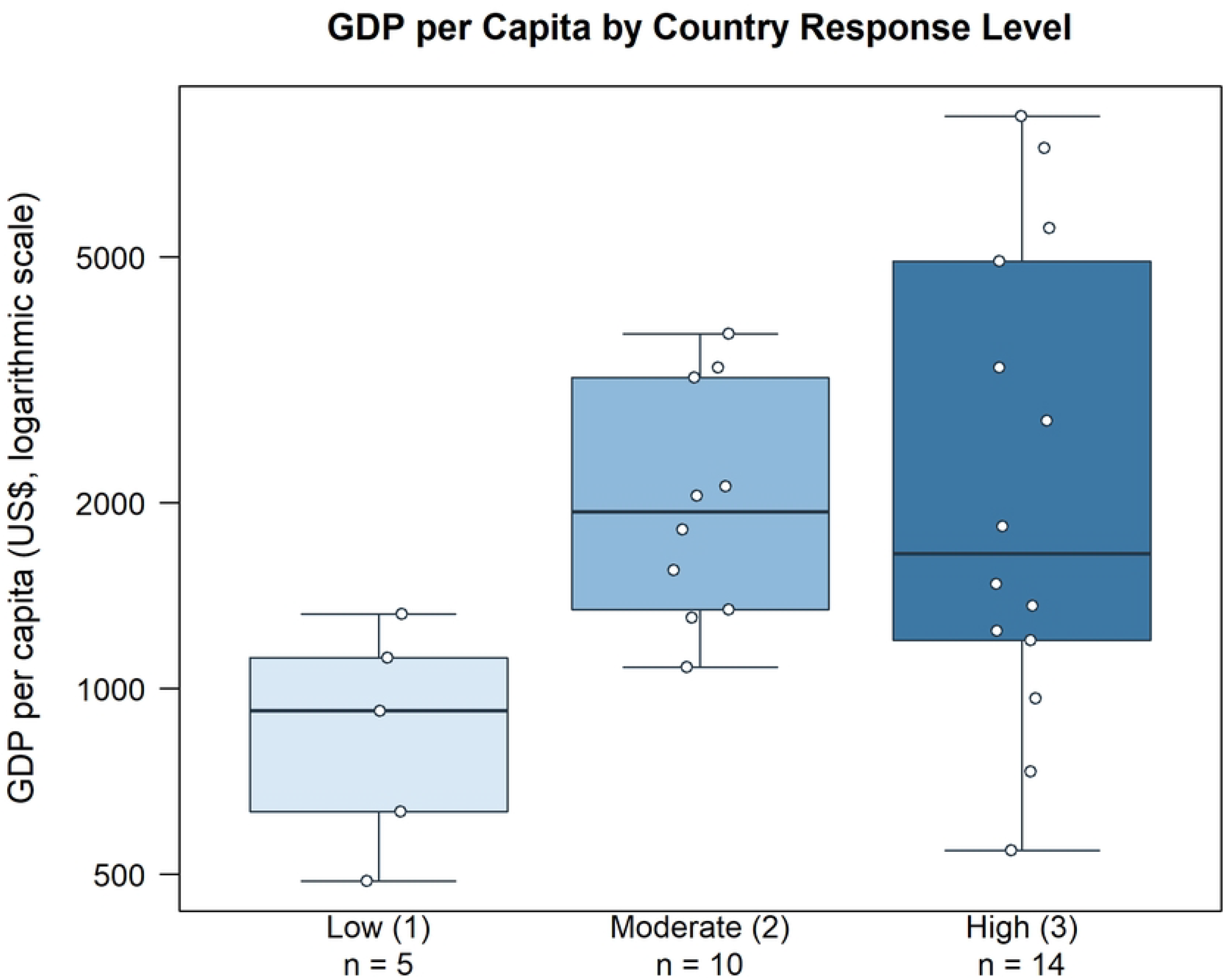
Distribution of Human Development Index by governmental response level.

**Fig 3.**
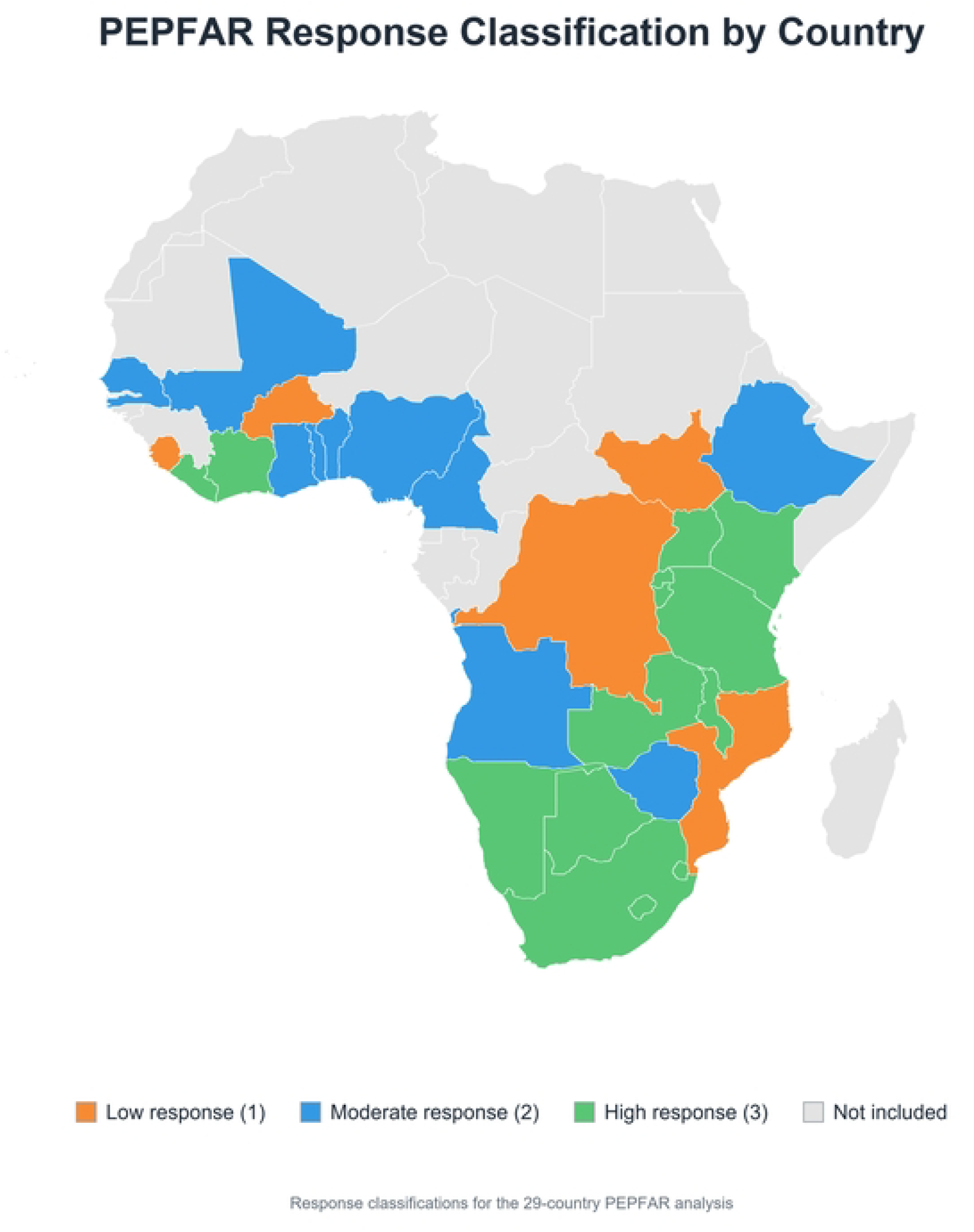
Distribution of GDP per capita by governmental response level.

Sensitivity analyses showed consistently positive associations for HDI and GDP per capita, although statistical significance varied across classifications. GDP per capita became significant in several scenarios, while HDI lost significance when South Africa was returned to its prior classification. All other factors remained nonsignificant. Using 2024 World Bank GDP per capita modestly attenuated the association with governmental response level (OR = 2.22; 95% CI, 0.94–5.24), without changing the primary conclusions. Complete regression results and sensitivity analyses are provided in **S2 Appendix**.

## Discussion

Higher-response countries were characterized by higher HDI. GDP per capita was also positively associated with response level in the unadjusted analysis, although this association did not remain significant after Holm correction. Historical PEPFAR funding demonstrated a positive but statistically nonsignificant association. These findings suggest that countries with greater human development and economic resources may have been better positioned to respond to abrupt funding disruptions, potentially reflecting stronger institutional capacity, healthcare systems, and program infrastructure.[3, 4, 14] However, given the ecological and cross-sectional study design, these associations should not be interpreted as evidence that higher HDI directly caused stronger country responses. Furthermore, because each predictor was evaluated separately, additional research is needed to distinguish the contributions of national development, economic resources, and historical PEPFAR investment to governmental response capacity.

Conversely, several Level 1 countries were experiencing active or recent conflict, including high HIV-burden nations such as Mozambique and the Democratic Republic of the Congo (DRC).[15, 16] These settings are of particular concern because healthcare delivery networks require years of investment and coordination to establish, but may deteriorate rapidly following funding interruptions.[3, 9, 17] Countries experiencing both conflict and substantial HIV burden may therefore be especially vulnerable to future increases in HIV transmission and mortality.[3, 4] Although many PEPFAR-supported activities have since resumed, the full impact of the 2025 funding disruptions may not become apparent for several years.[3, 4, 18] Furthermore, persistent limitations in mortality surveillance and HIV reporting systems may complicate efforts to accurately quantify their long-term effects.[19, 20]

An additional concern is that funding disruptions may disproportionately affect countries and populations already facing structural disadvantage.[1, 12] Several Level 1 countries have limited domestic resources available to replace external HIV funding, increasing the risk of treatment interruptions, workforce attrition, and supply chain instability.[1, 3, 9] Consequently, abrupt reductions in international HIV/AIDS funding have the potential to widen existing global health inequities and disproportionately affect populations with the fewest available alternatives for care.[1, 3, 9]

Five countries, Burkina Faso, the Democratic Republic of the Congo, Mozambique, Sierra Leone, and South Sudan, were classified as Level 1 response countries. These nations may represent priority targets for future stabilization efforts and investments aimed at preventing healthcare network deterioration following funding shocks. Although some Level 3 countries have larger absolute HIV burdens, they announced measures intended to maintain treatment infrastructure and mobilize alternative funding mechanisms. Several also announced plans to reduce long-term dependence on United States HIV/AIDS funding.[21, 22] While greater national self-sufficiency may ultimately strengthen health system resilience, the transition toward more autonomous financing structures has occurred under conditions associated with substantial disruption to existing HIV prevention and treatment programs.[3, 4, 12]

The primary limitation of this study is the classification framework and the inherent subjectivity involved in assigning governmental response levels. Different reviewers may employ different search strategies, identify different sources, or interpret the same information differently. These discrepancies do not necessarily indicate that any individual assessment was incorrect, but rather highlight the challenges of reducing complex governmental responses and healthcare systems developed over many years into a single categorical rating. Even when applying the same predefined criteria, differences in the available evidence and its interpretation may lead to varying assessments of the same country’s response. Although disagreements were addressed through additional review and consensus discussions, some degree of subjectivity remains unavoidable. **S1 Appendix** provides the individual classifications assigned by the three original reviewers, along with the fourth reviewer’s assessments of selected countries, to provide transparency regarding the variation in ratings and the subsequent adjudication process.

This study has several important additional limitations. First, country response classifications were derived from publicly available government announcements and third-party news reporting, which may incompletely capture internal governmental planning, informal mitigation strategies, or unpublished policy responses. Second, the rapidly evolving nature of PEPFAR-related policy changes means that national responses may continue to change after the study period, limiting the ability to fully characterize long-term governmental adaptation strategies. Third, this study evaluated response capacity rather than directly measuring downstream clinical outcomes such as HIV transmission, treatment interruption rates, or mortality changes, many of which may not become fully apparent for several years. Additionally, many high-burden countries continue to experience incomplete mortality surveillance and limited civil registration infrastructure, potentially impairing the accurate measurement of the true effects of funding disruptions.[19, 20] Finally, associations identified between response level and national indicators such as HDI or GDP per Capita should not be interpreted as causal relationships, but rather as exploratory findings intended to identify structural factors potentially associated with national HIV/AIDS system resilience.

## Conclusion

Reductions in foreign HIV/AIDS assistance are unlikely to affect all countries equally. Following a sudden external HIV financing shock when 2025 PEPFAR funding was disrupted, countries with greater underlying socioeconomic and institutional capacity were more likely to demonstrate substantial governmental mitigation responses. In addition, several Level 1 response countries were experiencing active conflict or political instability, further limiting their ability to sustain HIV prevention and treatment services during periods of funding uncertainty.[3, 4]

As HIV service delivery systems adapt to future funding challenges, efforts to improve resilience, diversify funding sources, and strengthen domestic capacity may help reduce vulnerability to abrupt external funding disruptions. Particular attention should be directed toward the five Level 1 response countries, Burkina Faso, the Democratic Republic of the Congo, Mozambique, Sierra Leone, and South Sudan, which demonstrated the lowest levels of governmental response following the funding freezes. The 2026 Ebola outbreak in the DRC provides a relevant example of how subsequent health emergencies can compound these vulnerabilities, placing additional pressure on health systems and disrupting access to HIV services.[23]

The long-term consequences of the 2025 PEPFAR and USAID funding interruptions remain uncertain and may not be fully understood for several years.[3, 4] However, the findings of this study provide an initial framework for identifying countries that may be at greatest risk during future funding shocks and may help guide efforts to strengthen HIV treatment and prevention infrastructure in settings with the greatest vulnerability.

## Data Availability

All data underlying the findings of this study are available within the manuscript and its Supporting Information files. Table 1 provides the country-level variables and final governmental response classifications. S1 Appendix contains individual reviewer assessments, consensus classifications, and supporting evidence, while S2 Appendix provides complete regression results and sensitivity analyses. The original publicly available country-level data were obtained from UNAIDS (https://aidsinfo.unaids.org/), the United Nations Development Programme (https://hdr.undp.org/data-center), the International Monetary Fund (https://www.imf.org/external/datamapper/datasets/WEO), PEPFAR (https://www.state.gov/pepfar/), and the World Bank (https://data.worldbank.org/indicator/NY.GDP.PCAP.CD).

## Acknowledgments

None

## S1 Appendix.

Individual reviewer classifications, adjudication results, and supporting evidence for governmental responses to the 2025 PEPFAR funding freeze across 29 Sub-Saharan African countries.

## S2 Appendix.

Univariable ordinal logistic and linear regression results and classification sensitivity analyses for governmental responses to the 2025 PEPFAR funding freeze.

## References

[1] Kates J, Wexler A, Lief E. Financing the Response to HIV in Low- and Middle-Income Countries: International Assistance from Donor Governments in 2015, https://files.kff.org/attachment/Financing-the-Response-to-HIV-in-Low-and-Middle-Income-Countries-International-Assistance-from-Donor-Governments-in-2015 (2016).

[2] PEPFAR’s Five-year Strategy. PEPFAR, https://2021-2025.state.gov/pepfar-five-year-strategy-2022/ (2022, accessed 10 June 2026).

[3] Stover J, Sonneveldt E, Tam Y, et al. Effects of reductions in US foreign assistance on HIV, tuberculosis, family planning, and maternal and child health: a modelling study. The Lancet Global Health 2025; 13: e1669–e1680.

[4] Hontelez JAC, Goymann H, Berhane Y, et al. The impact of the PEPFAR funding freeze on HIV deaths and infections: a mathematical modelling study of seven countries in sub-Saharan Africa. eClinicalMedicine 2025; 83: 103233.

[5] Reevaluating And Realigning United States Foreign Aid, https://www.whitehouse.gov/presidential-actions/2025/01/reevaluating-and-realigning-united-states-foreign-aid/ (2025, accessed 10 June 2026).

[6] Emergency Humanitarian Waiver to Foreign Assistance Pause. United States Department of State, https://www.state.gov/emergency-humanitarian-waiver-to-foreign-assistance-pause/ (2025, accessed 10 June 2026).

[7] Meyer-Rath G, Jamieson L, Mudimu E, et al. The cost of the plunge: the impact and cost of a cessation of PEPFAR-supported services in South Africa. AIDS 2025; 39: 1476–1480.

[8] Ratevosian J, Ngangula P, Tram KH. Modeling the fallout: projecting the global impact of donor funding cuts on HIV prevention, treatment, and care. Current Opinion in HIV and AIDS 2025; 20: 621–631.

[9] Jewell BL, Mudimu E, Stover J, et al. Potential effects of disruption to HIV programmes in sub-Saharan Africa caused by COVID-19: results from multiple mathematical models. The Lancet HIV 2020; 7: e629–e640.

[10] Impact of PEPFAR pause in South Africa, https://www.unaids.org/sites/default/files/20250224_impact-of-PEPFAR-pause-in-South-Africa.pdf (2025).

[11] Maintaining Essential Health Services and Systems, https://www.who.int/emergencies/diseases/novel-coronavirus-2019/technical-guidance/maintaining-essential-health-services-and-systems (accessed 10 June 2026).

[12] The Urgency of Now: AIDS at a Crossroads — 2024 Global AIDS Update, https://www.unaids.org/sites/default/files/media_asset/2024-unaids-global-aids-update_en.pdf (2024).

[13] Latest Global Program Results, https://2021-2025.state.gov/wp-content/uploads/2024/11/PEPFAR-Program-Results-Fact-Sheet-December-2024.pdf (2024).

[14] Bendavid E, Holmes C, Bhattacharya J, et al. HIV Development Assistance and Adult Mortality in Africa. JAMA 2012; 307: 2060–2067.

[15] Conflict in the Democratic Republic of Congo. Global Conflict Tracker, https://www.cfr.org/global-conflict-tracker/conflict/violence-democratic-republic-congo (accessed 10 June 2026).

[16] Mozambique | International Crisis Group, https://www.crisisgroup.org/africa/east-and-southern-africa/mozambique (2024, accessed 10 June 2026).

[17] Filiatreau LM, Musiello F, Tsagli DD, et al. PEPFAR interrupted: real-world consequences of U.S foreign aid instability for HIV service delivery in South Africa. The Lancet Regional Health -Africa 2026; 100039.

[18] PEPFAR Data Release. United States Department of State, https://www.state.gov/releases/office-of-the-spokesperson/2026/04/pepfar-data-release-2/ (accessed 10 June 2026).

[19] Kyu HH, Jahagirdar D, Cunningham M, et al. Accounting for misclassified and unknown cause of death data in vital registration systems for estimating trends in HIV mortality. J Int AIDS Soc 2021; 24: e25791.

[20] Mikkelsen L, Phillips DE, AbouZahr C, et al. A global assessment of civil registration and vital statistics systems: monitoring data quality and progress. The Lancet 2015; 386: 1395–1406.

[21] Rigby J, Mfula C, Kumwenda-Mtambo O, et al. Zambia pushed back on U.S. health funding deal to protect interests, government says. Reuters, 25 February 2026, https://www.reuters.com/sustainability/boards-policy-regulation/zambia-pushed-back-us-health-funding-deal-protect-interests-government-says-2026-02-25/ (25 February 2026, accessed 10 June 2026).

[22] Hingorani M. Global Health Watch: Zimbabwe steps away from US health MoU, new domestic funding in SA, LEN launched in Kenya + Zim, investment case for Global Fund. AVAC, https://avac.org/blog/global-health-watch-issue57/ (2026, accessed 10 June 2026).

[23] UNAIDS supports efforts to ramp up Ebola and HIV response amid evolving Ebola situation in the Democratic Republic of the Congo and Uganda | UNAIDS, https://unaids.org/en/resources/presscentre/pressreleaseandstatementarchive/2026/may/20260603_PR_Ebola_HIV_DRC (accessed 1 September 2026).

